# Real-time assessment of daytime sleepiness in drivers with multiple sclerosis

**DOI:** 10.1101/2020.06.19.20136077

**Authors:** Hannes Devos, Nesreen Alissa, Sharon Lynch, Maryam Sadeghi, Abiodun E. Akinwuntan, Catherine Siengsukon

**Affiliations:** Department of Physical Therapy and Rehabilitation Science, School of Health Professions, University of Kansas Medical Center, Kansas City KS; Department of Physical Therapy and Rehabilitation Science, School of Medicine, University of Maryland Baltimore, Baltimore MD; Department of Neurology, School of Medicine, University of Kansas Medical Center, Kansas City KS; Department of Exercise Physiology, Faculty of Physical Education & Sport Sciences, University of Tehran, Tehran, Iran; Office of the Dean, School of Health Professions, University of Kansas Medical Center, Kansas City KS

**Keywords:** multiple sclerosis, daytime sleepiness, PERCLOS, virtual reality, driving simulator

## Abstract

**Background:** Daytime sleepiness is a common symptom of multiple sclerosis (MS) that may jeopardize safe driving. The aim of this study was to compare daytime sleepiness, recorded in real-time through eyelid tracking, in a simulated drive between individuals with MS (iwMS) and healthy controls.

**Methods:** Fifteen iwMS (age = median (Q1 – Q3), 55 (50 – 55); EDSS = 2.5 (2 – 3.5); 12 (80%) female) were matched for age, sex, education, and cognitive status with 15 controls. All participants completed self-reported fatigue and sleepiness scales including the Modified Fatigue Impact Scale (MFIS), Pittsburg Sleep Quality Inventory (PSQI), and Epworth Sleepiness Scale (ESS). Percentage of eyelid closure (PERCLOS) was extracted from a remote eye tracker while completing a simulated drive of 25 minutes.

**Results:** Although iwMS reported more symptoms of fatigue (MFIS, p =0.003) and poorer sleep quality (PSQI, p = 0.008), they did not report more daytime sleepiness (ESS, p = 0.45). Likewise, there were no differences between groups in real-time daytime sleepiness, indexed by PERCLOS (p = 0.82). Both groups exhibited more real-time daytime sleepiness as they progressed through the drive (time effect, p< 0.0001). The interaction effect of group*time (p = 0.05) demonstrated exacerbated symptoms of daytime sleepiness towards the end of the drive in iwMS compared to controls.

PERCLOS correlated strongly (Spearman ρ = 0.76, p = 0.001) with distance out of lane in iwMS.

**Conclusion:** IwMS show exacerbated symptoms of daytime sleepiness during monotonous, simulate drive. Future studies are warranted to investigate the effect of MS on daytime sleepiness during real-world driving.

## 1. Introduction

Although fatigue and daytime sleepiness are sometimes used interchangeably, they encompass distinct constructs that are often affected in multiple sclerosis (MS).^1^ Both can be a consequence of sleep disorders.^2,3^ Fatigue is a “subjective lack of physical and/or mental energy that is perceived by the individual or the caregiver to interfere with usual and desired activities.”^4^ Fatigue is reported in up to 90% of individuals with MS (iwMS),^5^ and 40% of iwMS reports fatigue is their worst symptom.^6^ Although daytime sleepiness is less prevalent and severe than fatigue, a substantial percentage (19% to 53%) of iwMS report feeling sleepy or drowsy during the day.^3^ Daytime sleepiness is defined as the “inability to stay awake and alert during the major waking episodes of the day, resulting in periods of irrepressible need for sleep or unintended lapses into drowsiness or sleep.^7^ Daytime sleepiness is due to an imbalance in the sleep promoting and wake promoting neuronal systems.^1^

The occurrence of daytime sleepiness in MS gives rise to several concerns regarding its impact on specific aspects of daily life including driving. The lack of cognitive stimulation from monotonous driving may exacerbate daytime sleepiness. Lapses of drowsiness or sleep are associated with loss of vehicle control, and out-of-lane excursions.^8^ This puts drivers at an increased risk of motor vehicle crashes (MVC).^9^ Around 77% of iwMS continue driving after being diagnosed.^10^ Individuals with MS tend to alter their driving habits by driving less frequently and adopting more self-limiting behaviors.^11^ Although the vast majority of drivers with MS are fit to drive following an on-road test,^12^ they are reported to have greater frequency of driving violations,^13^ vehicle collisions,^14^ and hospital visits following a car crash^15^ compared to healthy controls. Impairments in cognitive functions, particularly in speed of processing, attention, and visuospatial and executive functions, have emerged as most important predictors of on-road driving performance.^16^ The effect of daytime sleepiness on driving performance in iwMS has yet to be established.

The Epworth Sleepiness Scale (ESS) is perhaps the most ubiquitous daytime sleepiness assessment of iwMS.^17^ Although the ESS provides a fast, reliable, and accurate assessment of daytime sleepiness, the questionnaire relies on subjective self-recall of the participant,^17^ and lacks the ability to monitor sleepiness continuously in real-life activities.^18,19^ Sleepiness is a gradual process that includes a sequence of physiological and behavioral changes.^20^ Observable changes in eye movement, eyelid behavior, head nodding, and facial expression have been associated with increased daytime sleepiness.^21^ The PERcentage of eye lid CLOSure (PERCLOS) on the pupil over time may be the most widely accepted eye tracking method for vigilance and sleepiness detection in human-machine interface studies, such as driving,^22^ and aviation.^23^ Unlike ESS, PERCLOS can be used to monitor daytime sleepiness in real-time during functional activities such as driving in iwMS.

The aim of this study was to compare the effect of a monotonous driving task on daytime sleepiness, indexed by PERCLOS, between iwMS and healthy controls.

## 2. Materials and Methods

### 2.1 Participants

Participants with a clinical diagnosis of MS according to the McDonald Criteria were recruited from the University of Kansas Multiple Sclerosis Clinic. Inclusion criteria were (1) between the ages of 18 and 65; (2) ability to understand the instructions in English; (3) in possession of a valid driver’s license; and (4) actively driving at the moment of testing with or without adaptive devices. Exclusion criteria were: (1) ocular motility problems such as nystagmus or cranial nerve palsy (III, IV, VI), (2) unresolved retina or pupillary conditions; (3) currently taking steroids, benzodiazepines, or neuroleptics; (4) exacerbations in the month preceding testing; (5) history of any substance abuse; and (6) history of a neurological disorder other than MS. From 06/01/2018 – 05/31/2019, 15 participants with MS were recruited and matched with 15 healthy controls according to age, sex, education, and cognitive status.

### 2.2 Procedure

All testing took place in less than 1.5 hours including consent and rest breaks. The study was approved by the University of Kansas Institutional Review Board. All participants provided informed consent.

#### 2.2.1 Demographic and clinical information

Age, sex, and education level were recorded. Cognition was evaluated using the Montreal Cognitive Assessment (MOCA).^24^ Clinical information included years since diagnosis, Expanded Disability Status Scale (EDSS) score,^25^ and type of MS. We also administered the Modified Fatigue Impact Scale (MFIS),^26^ the Pittsburg Sleep Quality Index (PSQI), and the Epworth Sleepiness Scale (ESS). The MFIS^27^ consists of twenty-one items divided into three components: physical, cognitive, and psychosocial. Each item is scored from 0 (never) to 4 (almost always) for a score ranging from 0–84. A higher score indicates a greater level of fatigue. The PSQI^28^ consists of 19 self-rated questions which form a global score ranging from of 0 – 21 with a higher score indicating poorer sleep quality. The ESS^29^ consists of 8 different scenarios of daily activities, and participants rate how likely they would be to fall asleep during each activity with 0 (no chance of dozing) to 3 (high chance of dozing). The global score ranges from 0 – 24 with a higher score indicates worse daytime sleepiness. Participants scoring 11 or higher on the ESS were considered having symptoms of daytime sleepiness.

#### 2.2.2 Driving simulation protocol

Daytime sleepiness was assessed using a house-modified virtual reality portable driving simulator (PDS) that was powered on STISIM Drive® (version 3, STI Inc, Hawthorne, CA) software (Figure 1). Images of the traffic scenario were projected on a single 22-inch screen with 45 degrees field of view. Participants used a Logitech Steering wheel and pedals to navigate through the scenario. The vehicle engine sound and ambient traffic noise were heard through the simulator loudspeakers. All instructions were recorded in the software program and automatically played as participants progressed through the scenario. The scenario started with a warm-up section of about five minutes (11,000 ft or 3,353 m) to gradually familiarize with the simulator software and hardware. In the familiarization, participants started on a two-lane road and were instructed to gradually increase their speed to 45 miles per hour. The two-lane road then transitioned into a four-lane road where participants were familiarized with highway driving by following a lead vehicle driving at a constant speed of 60 miles per hour. After the lead vehicle disappeared, the actual daytime sleepiness scenario started. This section took about 20 min to complete and was 99,600 ft (30,358 m) long and consisted of rural interstate driving on straight roads with speed limits of 70 miles per hour with no to little ambient traffic. The driver’s view was obstructed with dense fog that limited visibility beyond 1,000 ft (305 m) ahead of the driver. The lack of cognitive stimulation by the driving simulation was purported to exacerbate daytime sleepiness.

**Figure 1.**
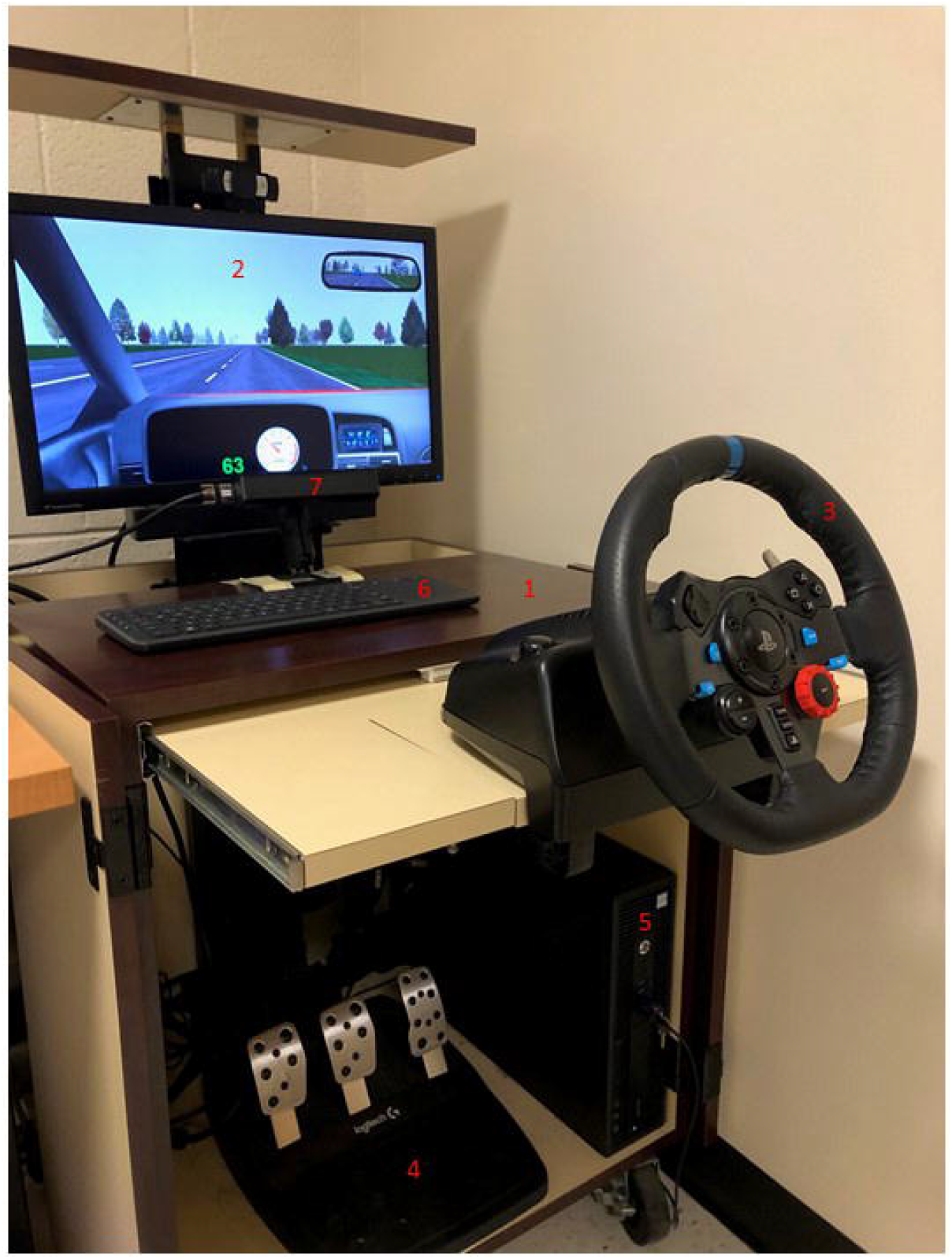
Portable Driving Simulator with FX 3 Eye Tracker 1. Shell 2. Simulator screen 3. Steering wheel 4. Pedals 5. Simulator computer 6. Keyboard 7. FX3 eye tracker

Following driving simulator parameters were recorded at 60 Hz: (1) Time to completion (s), (2) distance over speed limit (%), defined as the percentage of total distance drivers exceeded 70 mph; and (3) distance out of lane (%), defined as the percentage of total distance drivers crossed the center line or the road edge.

Any symptoms of driving simulator sickness (i.e., simulator adaptation syndrome or SAS), were monitored for during the drive. The Simulator Sickness Questionnaire (SSQ) was administered after the drive to assess SAS.^30^ None of the drives were aborted due to severe SAS.

#### 2.2.3 Eye lid closure recording

Eye lid closure was recorded while driving the simulator scenario using a remote eye tracker (FX3, SeeingMachines, Canberra, Australia) placed 11.4 cm in front of the screen with an upward angle of 12.8 degrees. Participants were seated 45 cm in front of the screen with their hands relaxed on the steering wheel.

The data extracted from the eye tracker and the driving simulator were synchronized using the Quad Server module of Eyeworks®. Eyelids were tracked using the EyeWorks® Facekit Module that recorded eyelid closure on a continuous scale ranging from 0 (eyes fully closed) to 1 (eyes fully open). PERCLOS was defined in our study as the percentage of a time interval that the eyes were 80% to 100% closed (exclusive of blinks).^31^ The total time (i.e. number of frames) of the daytime sleepiness scenario was divided into 10 equal parts. PERCLOS was extracted for each time epoch, i.e., 0 – 10; 11 – 20; 21 – 30; 31 – 40, 41 – 50, 51 – 60, 61 – 70, 71 – 80, 81 – 90, and 91 – 100 percent of completion time.

#### 2.2.4 Data analysis

Normality of variables was evaluated using visual inspection of histogram plots and Kolmogorov Smirnov tests. As can be expected due to relatively small sample size, assumptions of normality were violated in all continuous outcome measures, except for total scores on the MFIS. Consequently, non-parametric statistics were employed in all analyses. Group comparisons were evaluated using the Wilcoxon Rank Sum test for ordinal and ratio variables and the Fisher’s Exact test for nominal variables. Generalized linear mixed models with random intercept was employed to evaluate the main effects of group and time, and the interaction effect of group by time, on PERCLOS. Results of demographic clinical variables were correlated with the overall change in PERCLOS (last epoch – first epoch) using Spearman ρ correlations. All statistical analyses were conducted in SAS Enterprise Guide, version 8.2. P values ≤ 0.05 were considered significant.

## 3. Results

### 3.1 Demographic and clinical variables

We included 15 participants with MS and 15 controls matched for age, sex, education, and cognitive status (Table 1). All participants with MS had relapsing-remitting MS, except for one who was diagnosed with primary progressive MS. EDSS scores ranged from 1 to 6, indicating very mild to moderately advanced disease stage. However, the majority (75%) had mild to moderate symptoms of MS.

**Table 1.** Comparison of demographic, clinical, and driving performance variables between participants with MS and healthy controls

|  |  |  |  |
| --- | --- | --- | --- |
| Variable | MS (n = 15) | HC (n = 15) | P-value |
| *Demographic* |  |  |  |
| Age, years | 55 (50 – 59) | 48 (46 – 53) | 0.09* |
| Sex, female (%) | 12 (80) | 11 (73) | 0.31# |
| Education, years | 14 (12 – 16) | 16 (15 – 18) | 0.10* |
| *Clinical* |  |  |  |
| MoCA, total | 25 (23 – 27) | 27 (24 – 28) | 0.13* |
| Disease duration, years | 8 (4.5 – 15.5) | N/A |  |
| EDSS, total | 2.5 (2 – 3.25) | N/A |  |
| MFIS, total | 40 (27 – 54) | 15 (11 – 29) | 0.003* |
| MFIS, physical | 15 (11 – 25) | 5 (1 – 13) | 0.002* |
| MFIS, cognitive | 16 (12 – 31) | 13 (7 – 14) | 0.02* |
| MFIS, psychosocial | 2 (1 – 5) | 1 (0 – 3) | 0.12* |
| PSQI, total | 10 (6 – 13) | 4 (3 – 8) | 0.008* |
| ESS, total | 8 (5 – 13) | 8 (4 – 10) | 0.45* |
| *Driving* |  |  |  |
| Completion time, seconds | 1,153 (1,147 – 1,172) | 1,164 (1,145 – 1,196) | 0.46* |
| Distance over speed limit, % | 3.57 (0.27 – 10.30) | 1.65 (0 – 6.99) | 0.30* |
| Distance out of lane, % | 0.10 (0 – 0.29) | 0 (0 – 0.30) | 0.78* |
| Abbreviations: ESS, Epworth Sleepiness Scale; MoCA, Montreal Cognitive Assessment; MFIS, Modified Fatigue Impact Scale; PSQI, Pittsburgh Sleep Quality Index |  |  |  |
| *Wilcoxon Rank Sum test; #Fisher's Exact test. Variables are expressed as median (Q1 – Q3) or frequencies (%). |  |  |  |

**Table 2.** Spearman rho correlations between PERCLOS and demographic, clinical, and driving variables in the MS and control groups

| Variable | MS (n = 15) | P-value | HC (n = 15) | P-value |
| --- | --- | --- | --- | --- |
| *Demographic* |  |  |  |  |
| Age, years | 0.08 | 0.86 | -0.01 | .77 |
| Sex, female (%) | 0.14 | 0.61 | -0.12 | 0.65 |
| Education, years | -0.40 | 0.16 | -0.07 | 0.82 |
| *Clinical* |  |  |  |  |
| MoCA, total | -0.10 | 0.72 | 0.26 | 0.35 |
| Disease duration, years | 0.63 | 0.04 | N/A | N/A |
| EDSS, total | 0.35 | 0.20 | N/A | N/A |
| MFIS, total | -0.22 | 0.41 | 0.38 | 0.16 |
| PSQI, total | 0.09 | 0.74 | -0.10 | 0.71 |
| ESS, total | -0.21 | 0.44 | 0.006 | 0.98 |
| *Driving* |  |  |  |  |
| Completion time, seconds | 0.30 | 0.27 | 0.04 | 0.88 |
| Distance over speed limit, % | -0.06 | 0.83 | 0.30 | 0.28 |
| Distance out of lane, % | 0.75 | 0.002 | 0.29 | 0.28 |
| Abbreviations: ESS, Epworth Sleepiness Scale; MoCA, Montreal Cognitive Assessment; MFIS, Modified Fatigue Impact Scale; PSQI, Pittsburgh Sleep Quality Index |  |  |  |  |
| *Wilcoxon Rank Sum test; #Fisher's Exact test. Variables are expressed as median (Q1 – Q3) or frequencies (%). |  |  |  |  |

Participants with MS reported worse symptoms of fatigue than controls. The difference in total score on the MFIS (p = 0.003) was mainly driven by worse symptoms on the physical (p = 0.002) and cognitive subscales (p = 0.02) in iwMS. There were no group differences in the psychosocial subscale of the MFIS (p = 0.12).

Similarly, iwMS reported higher scores on the PSQI (p = 0.008), reflecting poorer sleep quality compared to healthy controls. By contrast, no differences were found in self-reported daytime sleepiness between the two groups (p = 0.45). Five (33%) iwMS reported ESS scores of 11 or higher while three (20%) in the control group reported scores higher than 11 (Fisher’s Exact, p = 0.68).

### 3.2 Driving simulator variables

None of the three driving simulator outcomes differentiated between the two groups (Table 1).

### 3.3 Percentage of eye closure over pupil

Figure 2 displays real-time daytime sleepiness, indexed by PERCLOS, in both groups as a function of percentage of completion time.

**Figure 2.**
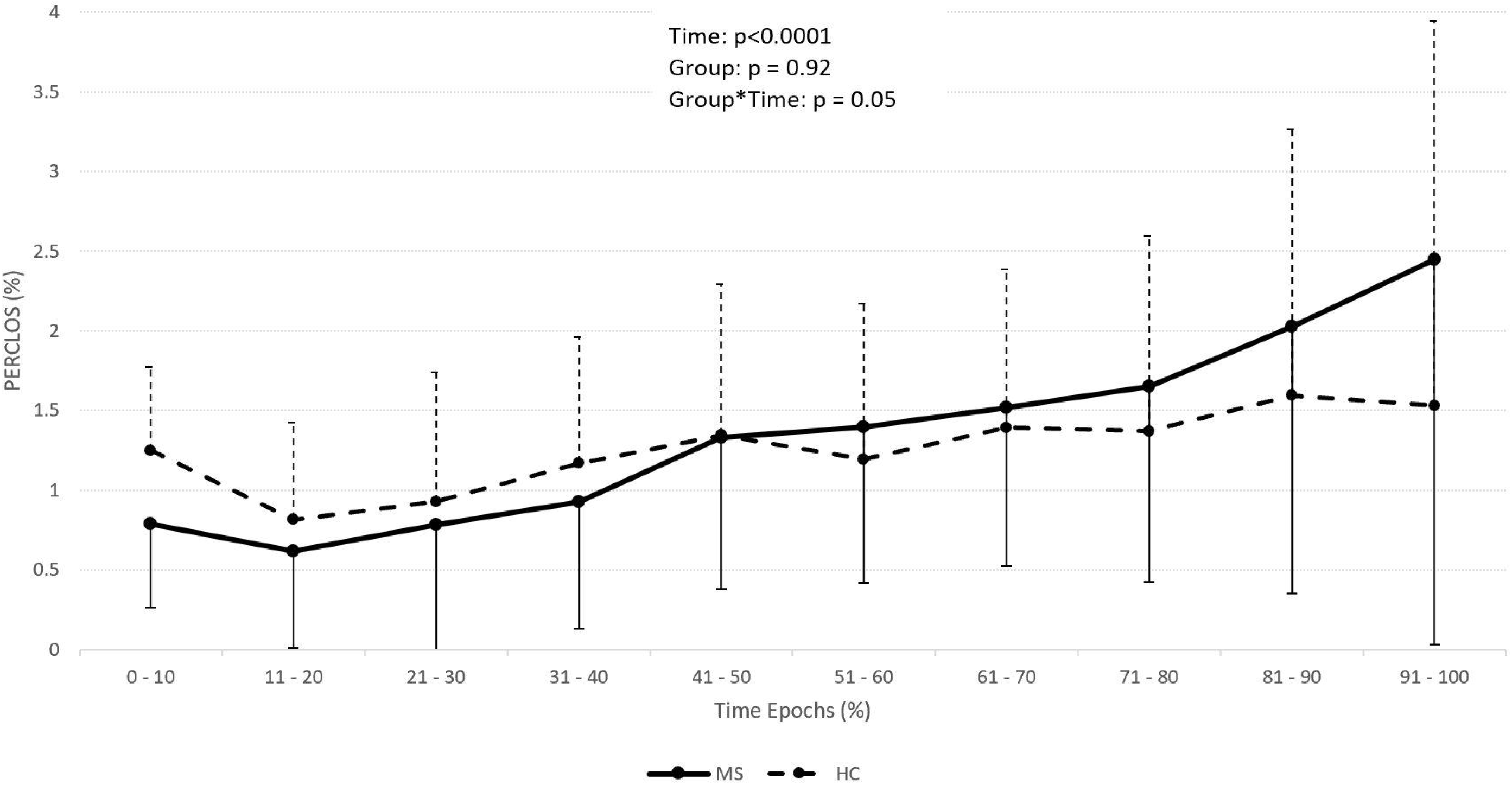
Comparison of PERCLOS as a function of completion time between drivers with multiple sclerosis (MS) and healthy controls (HC).

Generalized linear mixed modeling was employed to evaluate the effect of group, time, and the interaction effect of group by time on PERCLOS. Whereas group yielded no significant effect (p = 0.92), both groups showed more symptoms of daytime sleepiness as they progressed through the drive (p<0.0001). The interaction effect of group by time demonstrated that participants with MS showed a different pattern of PERCLOS throughout the duration of the test compared to controls (p = 0.05). Comparing the PERCLOS of last with first epoch, individuals with MS exhibited greater increase in daytime sleepiness (1.05% (0.40 – 2.14)) compared to healthy controls (0.31% (−0.40 – 1.19); p = 0.04). IwMS closed their eyes an average 2.5% of the time in the last epoch of the drive, whereas healthy controls only dozed off 1.5% of the time in the last epoch.

### 3.4 Correlations between demographic, clinical, driving simulator and change in PERCLOS

Whereas none of the demographic, clinical, or driving simulator variables correlated significantly with change in PERCLOS (last epoch – first epoch) in the control group, disease duration (ρ = 0.63, p = 0.04) and the percentage of distance out of lane (ρ = 0.76, p = 0.001) correlated strongly (> 0.50) with the change in PERCLOS in the MS group. No correlations were found between PERCLOS and any self-report daytime sleepiness, sleep quality, or fatigue scales.

## 4. Discussion

The aim of this study was to investigate the real-time effect of monotonous driving on daytime sleepiness in individuals with multiple sclerosis (iwMS). Our results showed that lack of cognitive stimulation from monotonous driving increased daytime sleepiness in both iwMS and controls. In addition, iwMS showed exacerbated symptoms of daytime sleepiness towards the end of the drive compared with controls. The lack of correlation between change in PERCLOS and ESS underscores the clinical importance of continuous, objective assessment in real-time during functional activities.

In a study by Neau et al., 32% of the iwMS had real-time assessed excessive daytime sleepiness, similar to the percentage (33%) found in our study.^32^ Although slightly higher, the prevalence of daytime sleepiness in iwMS did not differ statistically from that of control participants (20%). One study found iwMS had higher daytime sleepiness using the Pupillographic Sleepiness Test (PST) and lower general wakefulness using the computerized Test of Attentional Performance (TAP) than controls compared to normative data.^33^ Interestingly, iwMS without select comorbidities, such as anemia, thyroid dysfunction, depressive symptoms or antidepressants, had significantly higher daytime sleepiness on the PST compared to controls but iwMS with select comorbidities did not. Both MS groups also showed lower general wakefulness compared to the control group.^33^

By contrast, our results support previous studies that found no differences in self-reported and real-time daytime sleepiness between iwMS and controls. Scores on other real-time daytime sleepiness assessments, such as the Multiple Sleep Latency Test (MSLT), did not demonstrate differences between iwMS with fatigue, iwMS without fatigue, and controls. ^34^ Likewise, Taphoorn et al. reported no difference in the MSLT in a small sample of iwMS (n=16) selected due to “prominent” fatigue and sleep disturbances compared to controls.^35^ Neau et al. did not demonstrate differences in MSLT between iwMS who reported fatigue with or without self-report excessive daytime sleepiness, but there was no control group to serve as a comparison.^32^ Another study found no significant difference in real-time daytime sleepiness using pupillography between iwMS and controls.^36^ Our study was the first to assess real-time daytime sleepiness in iwMS using a prolonged driving task which may be a more ecologically valid assessment of daytime sleepiness than lying down or sitting in a quiet, dark room. Support of this assertation is that both groups showed more symptoms of daytime sleepiness as they progressed through the drive and iwMS exhibited a greater increase in daytime sleepiness despite the sample of iwMS and controls not reporting excessive daytime sleepiness (average ESS < 10 for both groups).

No significant correlations were found between self-reported symptoms of fatigue, sleep quality, or daytime sleepiness and PERCLOS measures. This is perhaps not surprising as these are likely associated albeit different constructs. Paucke et al. reported a significant correlation between real-time daytime sleepiness (PST) and self-report sleep quality (PSQI) only for iwMS with select comorbidities but not in controls or iwMS without select comorbidities.^33^ Furthermore, while iwMS without select comorbidities exhibited higher real-time daytime sleepiness than controls, there was no significant correlation with fatigue (MFIS) or sleep quality (PSQI). Furthermore, Frauscher et al. reported no correlations between real-time daytime sleepiness (pupillography) and ESS or Stanford Sleepiness Scale in the general sample (iwMS and controls) or in the iwMS group.^36^ In addition, Neau et al. reported MSLT was not correlated with ESS,^32^ and Kaynak et al. found no difference in ESS or MSLT between iwMS with and without fatigue, suggesting fatigue and daytime sleepiness may not be related.^34^ Furthermore, self-report measures can be influenced by social desirability, motives, memory, and depression whereas objective real-time assessments are likely less so.^33^

Whereas no significant? correlations between disease duration and self-reported daytime sleepiness were found in a previous study,^37^ our findings showed a significant correlation between disease duration and PERCLOS. Distance spent out of lane, an indicator of the ability to control the vehicle, has shown to correlate with daytime sleepiness in driving simulator studies.^38^ Both lateral vehicle control and daytime sleepiness are important risk factors of MVC. Up to 15% of all MVC are related to sleepiness^39^ and the risk of MVC when sleepy is between four and six times greater compared to driving while awake.^40^ Although the PERCLOS values remained low in both groups, the exacerbated symptoms of iwMS experienced at the end of the drive may increase the likelihood of MVC. This assumption should be tested in future studies.

The current study provides important knowledge on real-time daytime sleepiness in MS during functional activities. However, several limitations of the current study warrant caution in the interpretation of findings and generalization to the general population of drivers with MS. Our study sample was small and although we included participants with a wide range of disease severity, the majority had mild to moderate symptoms of MS. Likewise, only a minority of participants self-reported daytime sleepiness. Still, significant time and interaction effects were observed in PERCLOS in this group of participants with relatively preserved functions. We used a virtual reality driving simulator to mimic real-world driving. We cannot refute that driving behavior in the simulator may be different than driving in the real world. Although the scenario evoked daytime sleepiness, the drive was quite short (average of 25 minutes including warm-up). Since our findings showed a difference in PERCLOS toward the end of the drive, future studies should include longer drives to assess the effect of prolonged driving on daytime sleepiness in MS.

In conclusion, we established the importance of evaluating daytime sleepiness using objective, real-time measures in a functionally relevant context. Although no differences were found between iwMS and controls on PERCLOS, iwMS exhibited exacerbated symptoms of daytime sleepiness towards the end of the drive. Future research should include longer assessment of daytime sleepiness during real-world driving.

## Data Availability

Data are available upon request

## Acknowledgments

The authors thank Dr. Bunmi Morohunfola and Mr. Corey Gray for their assistance with data collection.

